# Feasibility study on using the diffusion-weighted imaging liver pulsation artifact as marker of liver fibrosis

**DOI:** 10.64898/2026.10.23.26358292

**Authors:** Fabian Groß, Guillaume Flé, Oliver Chaudry, Tri-Thien Nguyen, Islam Abbas, Sebastian Bickelhaupt, Sebastian Mathy, Carolin Arnold, Tobias Bäuerle, Hans-Peter Fautz, Konstantin Hellwig, Michael Uder, Marc Saake, Frederik B. Laun

## Abstract

**Background:** Liver diffusion-weighted imaging (DWI) often shows signal loss in the left lobe due to cardiac pulsation, known as the pulsation artifact.

**Purpose:** This study was conducted to turn this bug into a feature. We hypothesized that the pulsation artifact, caused by mechanically transmitted waves, is influenced by tissue stiffness and, thereby, indicative of cirrhosis in the left liver lobe. Thus, we evaluated the potential of DWI pulsation artifacts to detect liver cirrhosis.

**Study type:** This study included both retrospective patients and prospective healthy volunteers.

**Study population:** Thirteen healthy volunteers, 16 patients with cirrhosis (Ishak 5 or 6), and three patients without cirrhosis (Ishak 0, 1, or 2) were included.

**Field Strength/Sequence:** Imaging was performed at 1.5 T and 3 T using echoplanar DWI of the liver with two b-values, b50 = 50 s/mm² and b800 = 800 s/mm².

**Assessment:** After manual segmentation of the left and right liver lobes, 18 first-order features were extracted from b50 and b800 images. We assumed the pulsation artifact was predominantly present in the left liver lobe in the b800 images. Thus, we computed the ratios of the intensity-normalized b800 features obtained from the left and right liver lobes. Additionally, we performed an intensity normalization to address the sometimes different signal intensities of left and right liver lobe. Then, we conducted a receiver operating characteristic (ROC) analysis to differentiate cirrhotic from non-cirrhotic livers.

**Statistical tests:** ROC analysis and leave-one-out bootstrapping were applied, both yielding area under the curve (AUC) values.

**Results:** Left liver pulsation artifacts were more pronounced in non-cirrhotic livers and largely absent in cirrhotic livers. The first quartile feature, i.e. the first quartile of the normalized signal, best distinguished cirrhotic from non-cirrhotic livers (AUC = 0.95; 95% confidence interval 0.87 - 1.00).

**Data conclusion:** The left liver DWI pulsation artifact reflects liver stiffness and could serve as a simple, intrinsic marker of liver cirrhosis. Analysis of the signal magnitude can differentiate cirrhotic from non-cirrhotic livers.

## Introduction

Diffusion-weighted imaging (DWI) is a valuable tool for detecting and characterizing abdominal lesions (1,2). DWI was reported to be superior to standard T_2_-weighted sequences for the detection of small liver lesions (3–5). Usually, a small b-value (b < 150 s/mm²) and a high b-value (b > 500 s/mm²) are applied (6–9). The small-b-value image is used as an anatomical reference, since the signal loss due to diffusion weighting is negligibly small at this b-value. Application of small b-value diffusion encoding yields black-blood images due to the intravoxel incoherent motion (IVIM) effect, as the intravascular signal is suppressed (10). This provides an advantage for lesion detection over bright-blood T2-weighted imaging. The high-b-value image is used for lesion detection and characterization, as it enables computation of the apparent diffusion coefficient (ADC), which often differs across lesion types (6,7).

DWI sequences are sensitive to both diffusion and other types of motion, such as bulk, respiratory, or cardiac motion. In the liver, one challenge is the cardiac motion artifact (11–15) arising from a phase dispersion caused by cardiac pulsation. Pulsation waves propagate into the liver, leading to additional signal loss in diffusion-weighted images and resulting in ‘black holes’ on the images. The artifact is most pronounced in the left liver lobe due to its proximity to the heart (16,17). The artifact is worse at higher b-values (e.g., b >500 s/mm²) than at lower b-values (e.g., b <150 s/mm²) (18). Usually, this pulsation artifact is considered a nuisance. For example, it was the reason the left liver lobe was excluded from further analysis in previous studies (19,20) and has led to reduced sensitivity in detecting left-liver metastasis (16,21).

It seems logical to hypothesize that the severity of the pulsation artifact is related to the liver’s mechanical properties. If this were the case, this ‘bug’ (the pulsation artifact) could be turned into a feature (a proxy for liver stiffness and cirrhosis). Although liver biopsy is the standard for chronic liver disease diagnostics, this invasive procedure is limited by potential complications, such as pain, bleeding, sampling error, high cost, liver disease heterogeneity, patient reluctance, and a very small risk of death (22).

Magnetic resonance elastography (MRE) is a noninvasive approach for assessing stiffness (23–25). The standard MRE approach is to measure motion-weighted phase maps of the tissue while using an external actuator to generate tissue vibration. The phase maps are then converted into stiffness maps. Liver fibrosis is generally associated with hepatic tissue stiffening. Presumably for this reason, MRE is regarded as the most accurate noninvasive imaging technique for liver fibrosis evaluation (26,27). Indeed, MRE is recommended by multiple clinical societies, including the American College of Radiology (ACR) (28).

One limitation of liver MRE is that additional equipment is required, including an transducers that must be properly placed on the patient. To overcome this limitation, *intrinsic* MRE has been proposed, in which the liver’s natural motion is detected using, for example, motion-resolved tagging sequences (27,29). Although this approach is successful, it necessitates acquiring additional images, thereby prolonging the scan time.

This retrospective study was conducted to evaluate whether the DWI pulsation artifact can be used to detect liver cirrhosis. This approach would overcome the need for additional equipment in extrinsic MRE and the requirement to acquire extra sequences in the intrinsic MRE methods proposed so far.

## Material and Methods

### Retrospective data curation

The university’s IRB waived the need for informed consent for our retrospective data analysis (IRB number aaa, Ethics committee of the University bbb). The retrospective imaging data comprised datasets acquired during routine clinical practice at the host institute and were retrieved from the host institute’s Picture Archiving and Communication System (PACS) system from 08.01.2025 to 18.02.2025. A patient list for data extraction was obtained from the hospital information system via a search of the radiology information system for liver MRI scans performed between 2016 and 2024, both including cases with documented or suspected hepatocellular carcinoma (HCC), defined by histopathology or LI-RADS imaging criteria (30). Patients were only included when histopathological information, i.e. the Ishak score, was available.

Data quality was rated on a scale from 1 (best) to 5 (worst). The extent of cancerous tissue regions was rated on a scale from 1 (no cancer visible in the considered slice) to 5 (virtually no liver tissue identifiable in the relevant region). These ratings were performed by F.F. (final-year physics student), who was supervised by F.F.F. (physicist with >15 years of experience in abdominal MRI) for data quality assessment and T.T.T. (board-certified radiologist) for cancer extent assessment. Datasets with a score exceeding 3 were excluded.

### Prospective magnetic resonance imaging

Additionally, we acquired prospective data (approved by the local Institutional Ethics Committee, study number 24-381-Bm). Each participant provided written informed consent. Fifteen healthy volunteers were scanned on a clinical 1.5 T scanner (MAGNETOM Sola, Siemens Healthcare, Erlangen) using a vendor-provided 18-channel anterior body matrix coil in combination with a vendor provided 32-channel spine array coil from 17.04.2025 to 18.06.2025. The technical details of the acquisitions performed are listed in Table 1. Datasets were excluded when the data quality was rated as insufficient.

**Table 1.** MRI parameters. DWI = diffusion-weighted imaging, EPI = echo planar imaging, GRAPPA = GeneRalized Autocalibrating Partial Parallel Acquisition, SMS = simultaneous multislice, MDDW = multi-directional diffusion weighting, SPAIR = Spectral Adiabatic Inversion Recover.

| Sequence | DWI EPI (volunteers) | DWI EPI (patients) |
| --- | --- | --- |
| Repetition time (ms) | 3,400 | 1,900 - 2,500 (6900 once) |
| Echo time (ms) | 55 | 44 - 65 |
| Echo spacing (ms) | 0.49 | 0.54 |
| Voxel size (mm<sup>3</sup>) | 1.9 × 1.9 × 5 interpolated to 1.56 × 1.56 × 5 | (0.53 × 0.53 × 5) - (1.61 × 1.61 × 5) |
| Field of view (read × phase; mm<sup>2</sup>) | 432 × 326.25 | (143 × 143) - (380 × 420) |
| Phase direction | Anterior-posterior | Anterior-posterior |
| Phase resolution | 100 % | 100 % |
| b-values (s/mm<sup>2</sup>) | 50, 800 | 50, 800 |
| Averages (b50, b800) | 2, 4 | 1, 2 |
| Diffusion mode | MDDW with 6 directions | 3- or 4-Scan Trace |
| Diffusion scheme | Monopolar (= single refocused) | Monopolar (= single refocused) |
| Number of slices | 40 (axial) | 34 - 47 (axial) |
| Slice distance | 20 % | 20 % |
| Parallel imaging | GRAPPA with acceleration factor 2 | GRAPPA |
| Nominal acquisition time (min:s) | 2:32 | 1:00 - 2:35 |
| Bandwidth (Hz/pixel) | 2,350 | 2,332 - 2,488 |
| Fat saturation | SPAIR | SPAIR |
| Trigger | Resp. Trigger (expiration) | Resp. Trigger (expiration) or free breathing |
| Phase Partial Fourier | Off | Off |

### Segmentation

Segmentations were performed by F.F. using 3D Slicer (version 5.81), under the guidance and supervision of T.T.T. For each patient and volunteer, the left and right liver lobes were segmented (liver segments 1 - 4 and 5 - 8, respectively). The inferior vena cava and the middle hepatic vein were used as landmarks to ensure that the right and left liver lobes were divided at the correct boundary. The segmentations spared the well-delineated portions of the portal vein and the right hepatic vein. As the extent of the liver regions that are affected by cirrhosis is very difficult to determine, we did not try to differentiate cirrhotic from non-cirrhotic liver regions for the segmentation. All segmentations were performed on b = 50 s/mm² trace-weighted images. T_1_-and T_2_-weighted images were used as guidance in the segmentation process.

### First-order feature extraction

First-order radiomics features were computed in Python 3.13.6 for the segmentations (i.e., separately for the right and left liver lobes). In this process, NIfTI files of the trace-weighted images were loaded together with the corresponding segmentations. File handling was performed using the Python packages os and pandas, whereas the numpy and seaborn packages were used to calculate first-order statistical features.

We aimed to compare the features of the right and left liver lobes. Consequently, we performed the following computation for each feature:

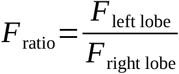

where *F*left lobe is the respective feature of the left liver lobe, and *F*right lobe that of the right liver lobe. This was performed once for the b = 50 s/mm² trace-weighted images and once for the b = 800 s/mm² trace-weighted images, yielding *F*ratio,b50 and *F*ratio,b800, respectively. As we observed global intensity variations in the left and right liver lobes, attributed to an incomplete suppression of surface coil flare, we additionally performed the following normalization step:

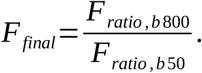

*F*final was used for the subsequent analysis.

Sixteen of the 19 standard first-order statistical features obtained from the radiomics data were used in the analysis. The radiomics features *range*, *minimum*, and *maximum* were excluded because we deemed them to be overly dependent on outliers. Two additional features were included: the first quartile (*q1*, the 25th percentile) and the third quartile (*q3*, the 75th percentile). Thus, 18 statistical features were employed for the subsequent analysis.

### Statistical analysis

Statistical analyses were performed in R (R Core Team, version 4.3.2). Given the small dataset, all analyses were performed with an explorative focus. We divided the datasets into two groups (group 1: healthy volunteers and Ishak score *≤* 2; group 2: Ishak score *≥* 5). The features were standardized using z-transformation before analysis.

First, a univariate analysis of the area under the curve (AUC) was conducted on the whole dataset using the individual features. The AUCs were computed with logistic regression models. The 95 % confidence intervals (CIs) of these univariate AUC values were computed with the function *ci.auc* in the R package pROC.

Second, a leave-one-out analysis was performed to minimize the overfitting risk inherent in the whole dataset analysis. This analysis was conducted once for the best-performing feature across the whole dataset. Additionally, this analysis was performed bivariately, combining the best-performing feature with any other feature. Since many features were highly correlated, we removed features with a Pearson correlation coefficient ≥0.8 with the best-performing feature in this bivariate analysis. For the remaining features, the predictive performance was estimated using an adaptation of the leave one out bootstrap (LOOBT) approach proposed in (31), in which the mean squared relative error was used as the performance metric instead of the AUC value (code provided as supplemental material). In this approach, logistic regression models with Firth penalization (*logisV* from the R package logistf) were fitted to stratified bootstrap samples (n = 2000). This approach was chosen because LOOBT provides approximately unbiased AUC estimates. CIs were also derived via bootstrapping according to the mean of out-of-bag predictions per patient.

## Results

### Study population

Twenty-two out of the 79 patients were excluded due to poor data quality or the presence of cancerous lesions in the right or left liver lobe that made the assessment of liver tissue unfeasible. Of the remaining 57 patients, 39 had cirrhotic livers, and 18 had non-cirrhotic livers. Biopsy results were available for 19 patients (16 cirrhotic and 3 non-cirrhotic). Among these, three patients had an Ishak score ≤ 2, four had a score of 5, and 12 had a score of 6. Table 2 summarizes details about the study population. The age of the remaining 19 patients was (67.8 ± 7.4) years (6 female, 13 male).

**Table 2.** Study population. The left column shows the Ishak scores of the patient data set. Right column represents the Siemens scanners used for acquisition.

| Patient Ishak score (0–6) | MRI system (patients) |
| --- | --- |
| 4–5 | Aera |
| 6 | Aera |
| 6 | Aera |
| 6 | Aera |
| 6 | Aera |
| 6 | Aera |
| 6 | Aera |
| 5 | Sola |
| 6 | Aera |
| 6 | Aera |
| 5 | Aera |
| 6 | Vida |
| 5 | Vida |
| 6 | Vida |
| 6 | Sola |
| 6 | Aera |
| 1 | Sola |
| 0 | Aera |
| 1 | Altea |

Two of the 15 healthy volunteer datasets were excluded due to poor data quality. The age of the remaining 13 volunteers was (26.5 ± 6.3) years (4 female, 9 male).

### Representative images

Figure 2 shows representative trace-weighted diffusion-weighted images acquired with a b-value of 800 s/mm². The slice number is indicated in the upper left corner of each image. The first column depicts a true positive cirrhotic case (Ishak score 6) without any visible pulsation artifacts across the slices. The second column illustrates a true negative healthy volunteer exhibiting visible signal voids (white arrows). Representative examples of a cirrhotic liver (Ishak score 5) exhibiting a pulsation artifact (false negative) and a non-cirrhotic liver (Ishak score 1) without visible artifacts (false positive) are shown in the appendix.

**Figure 1.**
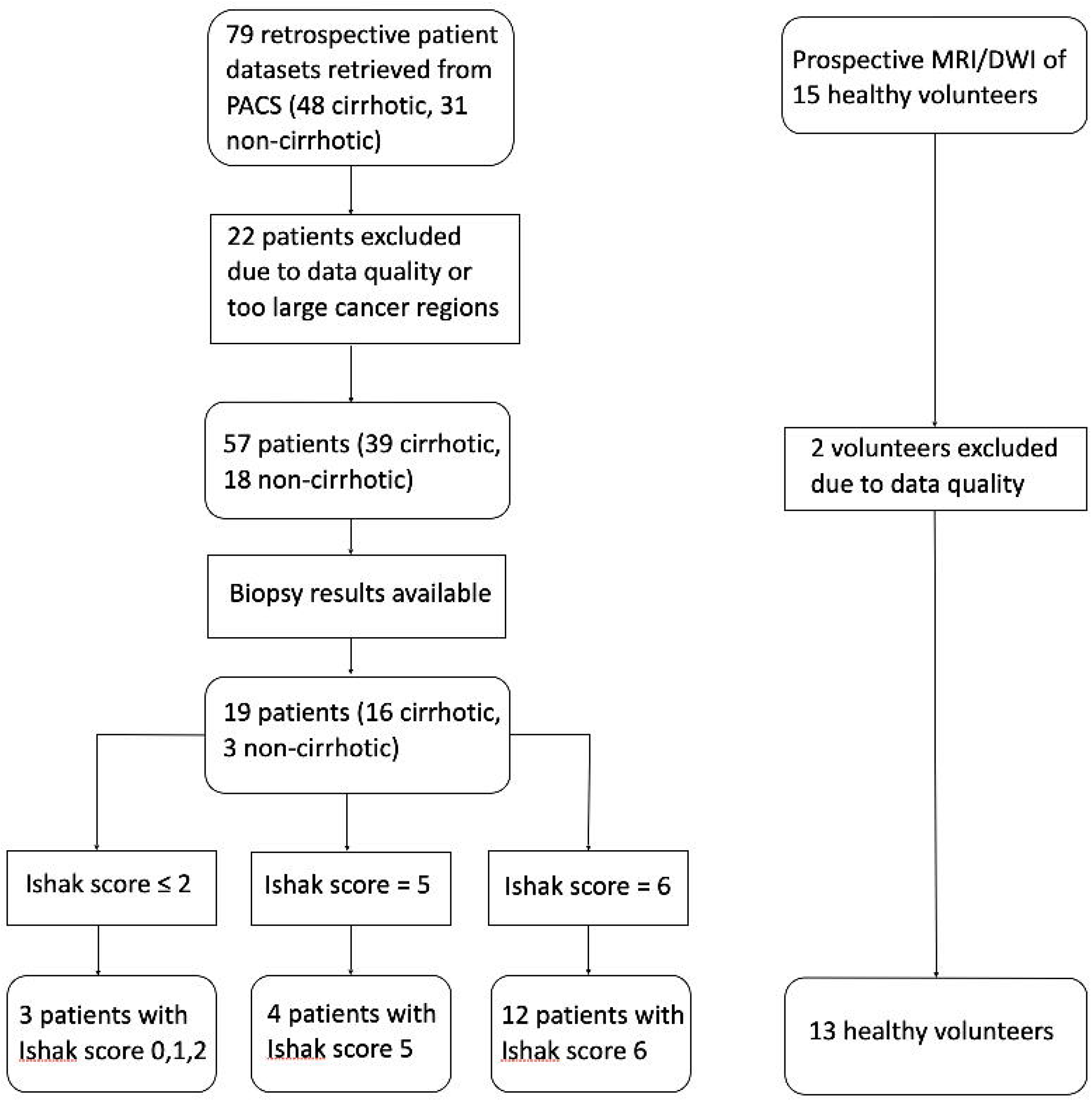
Study flowchart.

**Figure 2.**
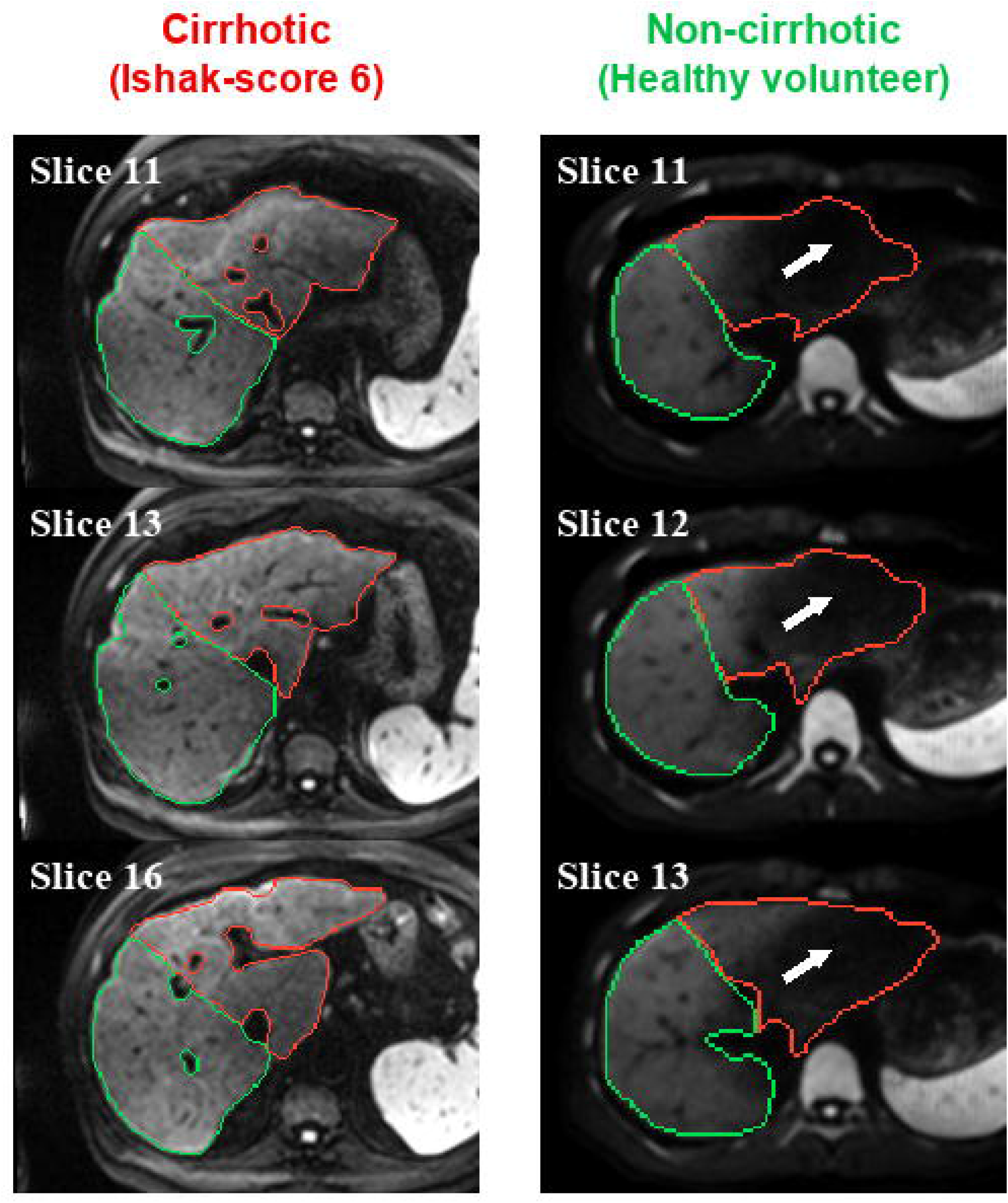
Representative examples of true-positive (cirrhotic, Ishak score 6) and true-negative (non-cirrhotic, healthy volunteer) cases. Each column shows three b = 800 s/mm² trace-weighted images of a cirrhotic or healthy patient. The left column shows a patient with cirrhosis and no visible artifact, the right column a healthy volunteer with an apparent artifact, indicated by white arrows. The green region marks the right liver lobe and the red region the left liver lobe. Arrow: Pulsation artifact.

### Quantitative analysis

Figure 3a shows boxplots of *q1*, which was the feature with the highest AUC in the whole-dataset analysis (AUC 0.95, CI: 0.87 - 1.00). Green data points represent healthy volunteers, yellow markers represent patients with Ishak scores of 0, 1, or 2, and red markers represent patients with Ishak scores of 5 (red crosses) or 6 (red dots). The *q1* values of cirrhotic livers were generally larger than those of non cirrhotic livers. Among the non-cirrhotic *q1* values, the Ishak 0–2 cases (orange dots) exhibited some of the largest values. These Ishak 0 - 2 *q1* values ranged from 0.71 to 0.83, thus mostly larger than the *q1* values of the healthy volunteers (green dots, *q1* = 0.40 to 0.68) and in the lower range of the cirrhotic cases (*q1* = 0.63 to 0.99).

**Figure 3.**
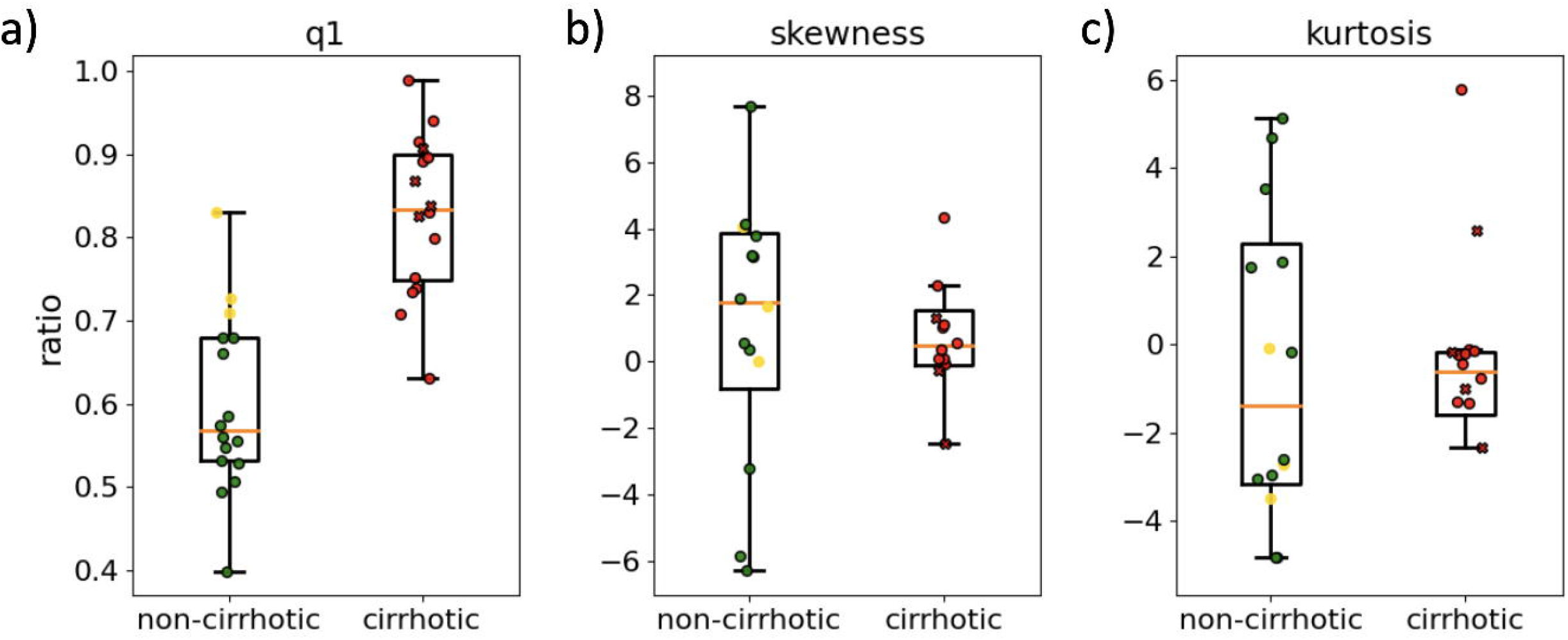
Boxplots for the features *q1*, *skewness*, and *kurtosis*. Green dots indicate healthy volunteers, orange dots represent patients with Ishak scores ≤2, red dots represent patients with Ishak scores of 6, and red crosses indicate patients with Ishak scores of 5. Outliers for *skewness* that are not shown: 335.5, -18,8 (healthy volunteers); -15.0, 366.7, - 9.62, 925.3 (Ishak score ≥5). Outliers for *kurtosis* that are not shown: 52.5, - 36.0 (healthy volunteers); - 62.9, - 44.6, - 1.9 (Ishak score ≥5).

Figure 3b shows the two features that led to the best AUC in the bivariate leave-one-out analysis (in combination with *q1*). Compared with *q1*, the overlap of the cirrhotic and non-cirrhotic cases increased substantially in *skewness* and *kurtosis*. Unlike in *q1*, the Ishak 0 - 2 values for these features (orange dots) appear randomly distributed across cases.

Figure 4a shows the AUC values obtained from the whole-dataset univariate analysis. The confidence intervals are shown as black bars. *q1* exhibited the highest AUC (0.95, CI: 0.87 - 1.00). Of the remaining features, the seven with the highest AUCs were highly correlated with *q1* (Pearson correlation coefficient ≥0.8, marked with red bars). These are mostly intensity-based features. The next two best performing features were *p90* (AUC 0.79, CI: 0.64 - 0.89), which is also an intensity based feature, and *std* (AUC 0.78, CI: 0.64 - 0.87). The features *skewness* and *kurtosis*, which performed best in the leave-one-out bivariate analysis in combination with *q1*, demonstrated low AUCs (0.56 with CI 0.35 - 0.77 and 0.54 with CI 0.32 - 0.76, respectively).

**Figure 4.**
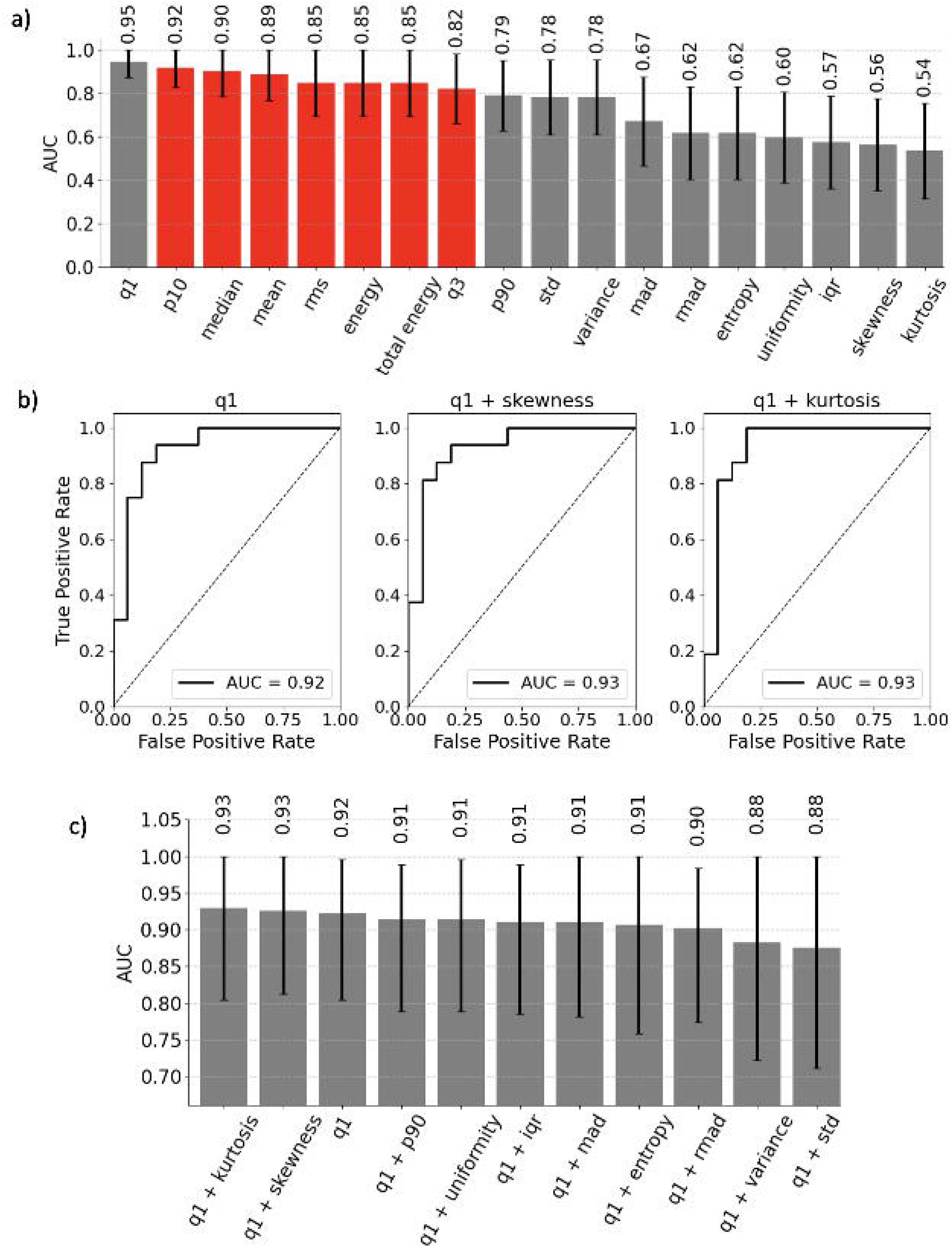
Receiver operator characteristics evaluation. a) Area under the curve (AUC) for all first-order features obtained in the whole-dataset analysis. Red bars show features with correlations larger than 0.8 compared with *q1*. Black bars represent the confidence intervals. b) ROC curves for the leave-one-out bootstrap (LOOBT) analysis: univariate *q1* and bivariate *q1* + *skewness*, *q1* + *kurtosis*. c) LOOBT analysis for the bivariate analyses of *q1* + features with correlations lower than 0.8 to *q1*. Additionally, the AUC for the univariate *q1* analysis is shown. Confidence intervals are shown as black bars.

Figure 4b shows the ROC curves for the leave-one-out analysis for the univariate feature *q1* and the bivariate feature combinations *q1*+*skewness* and *q1*+*kurtosis*. The ROC curves appear almost identical.

Figure 4c shows the AUC values obtained in the leave-one-out analysis. The AUC of the univariate feature *q1* was 0.92 (CI 0.80 - 1.00), smaller than that in the whole-dataset analysis shown in Fig. 4a.

The intention of the leave-one-out analysis is to avoid the overfitting that may occur in the whole dataset analysis. Naturally, this results in a tendency towards smaller AUC values. The feature combinations *q1*+*kurtosis* and *q1*+*skewness* yielded the highest AUC values in this analysis (AUC 0.93, CI: 0.80 - 1.00, and AUC 0.93, CI: 0.81 - 1.00, respectively). The combination *q1*+*std* resulted in the lowest AUC (0.88, CI 0.71 - 1.00).

Figure 5 shows the correlation matrix of all features. Strong positive correlations are depicted in red tones, whereas negative correlations are represented in blue tones. The numbers in each cell indicate the calculated correlation coefficients for each feature pair. A strong correlation among these signal intensity-related features was observed.

**Figure 5.**
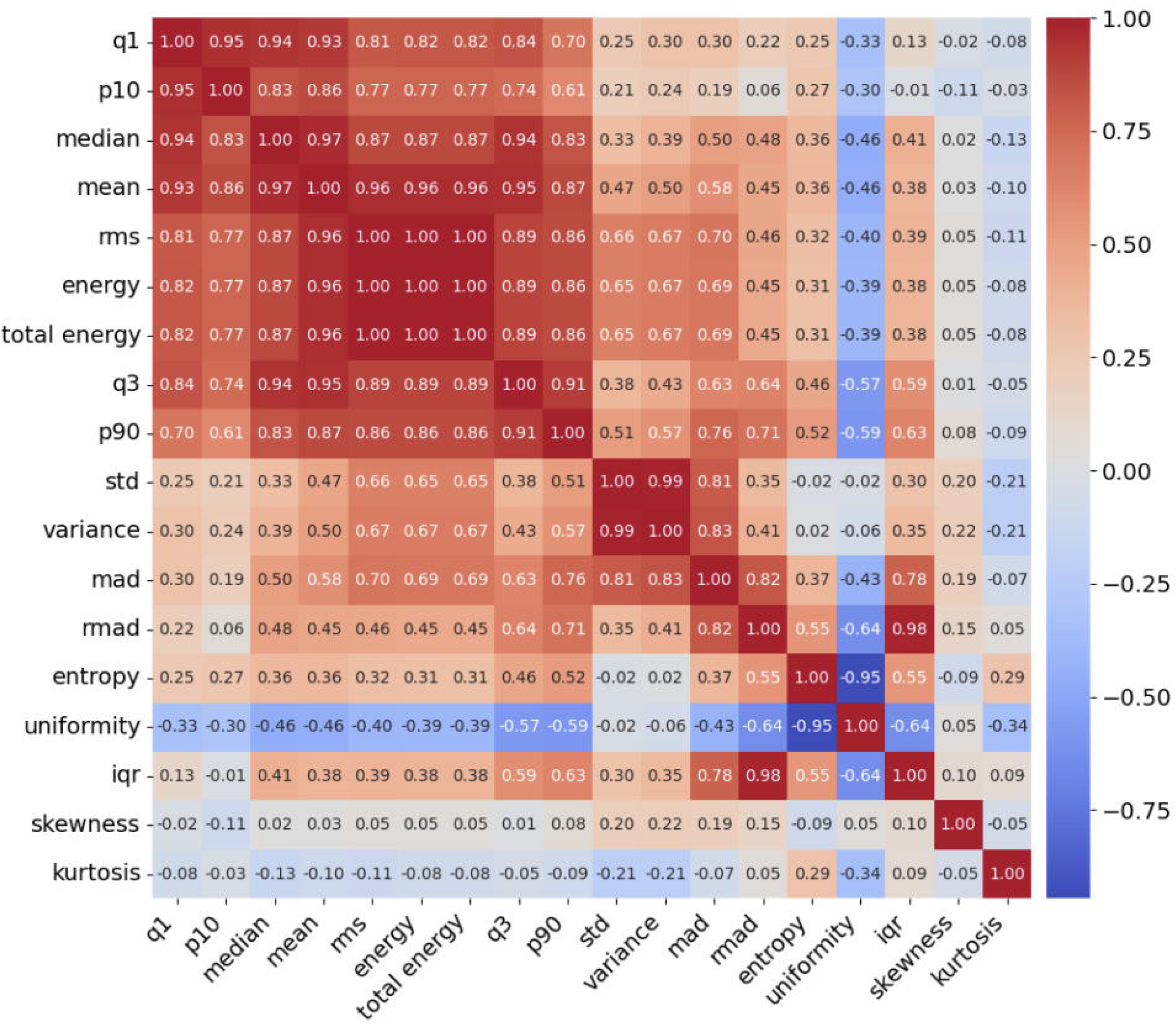
Correlation matrix for first-order features.

## Discussion

This study demonstrates that the DWI pulsation artifact, which generally appears most prominently in the left liver lobe and which is usually regarded as a nuisance, can serve as a potential indicator of liver cirrhosis. By quantitatively evaluating first-order statistical features derived from the left and right liver lobes, we distinguished cirrhotic from non-cirrhotic livers with high diagnostic accuracy.

From all evaluated parameters, the univariate *q1* feature performed best with an AUC of 0.93 in the leave-one-out analysis. The bivariate analysis of *q1* combined with *kurtosis* (or *skewness*) increased the AUC to 0.94; however, as *kurtosis* and *skewness* performed poorly in the univariate analysis (see Fig. 3), this effect appears to be a statistical artifact. Thus, using only one of the intensity-based features, such as the *mean* or *median*, which were all highly correlated to *q1* and exhibited similarly high AUC values, might be sufficient for detecting cirrhotic livers. The slightly better performance of *q1* might be explained by the fact that the pulsation artifact sometimes only appears in localized parts of the left liver lobe, which would be captured better by *q1* than by the median, for example. These findings support the hypothesis that the strength of the DWI pulsation artifact is related to the liver’s mechanical properties and could therefore serve as a simple, noninvasive marker of liver stiffness and cirrhosis.

The potential of DWI for diagnosing liver fibrosis has been evaluated previously. For example, Wang et al. demonstrated that MRE achieves better predictive accuracy than DWI for distinguishing different stages of liver fibrosis (32). However, this traditional approach focuses on measuring the ADC in artifact-free regions. For example, Wang et al. placed the ROIs in the right liver lobe, ensuring that “artifacts from motion and liver tissue with poor signal noise were carefully avoided”. Here, we propose an opposite approach: specifically examining the pulsation artifacts.

Other non-DWI imaging features have been associated with liver fibrosis but were found to be less suitable than MRE for this task (24). This makes sense, since these features are generally not directly linked to liver stiffness, unlike MRE and the DWI pulsation artifact.

Liver stiffness has been reported to increase progressively with increasing fibrosis grade. For example, Yin et al. reported that liver stiffness scales like an exponential function *≈* exp ( 0.24 ) *⋅ Fx*, where *Fx* = 0, 1, …, 4 (33). Thus, in future studies, it may be interesting to evaluate a functional relationship between the strength of the pulsation artifact and the fibrosis grade. Interestingly, Yin et al. reported an increase in liver stiffness from the normal liver to the F0 stage. This is consistent with our finding that the Ishak1 and Ishak2 cases exhibited higher *q1* values than healthy controls (see Fig. 3).

Our approach samples only the left liver lobe and may miss fibrotic changes that are present predominantly in the right liver lobe. In contrast, MRE provides maps of liver stiffness and thereby resolves liver stiffness heterogeneity. In principle, this is important as hepatic fibrosis can be heterogeneous throughout the liver (34) and MRE can provide guidance when performing biopsies (35). In practice, this advantage of MRE may not be overly relevant because MRE stiffness evaluations are usually performed using liver-averaged quantifications.

### Limitations

Our study has several limitations. The sample size is limited, and more reliable results could be obtained by including additional patient datasets. In addition, the retrospective cohort was limited to patients who underwent HCC-related MRI examinations, which can have led to a selection bias. A further limitation is that we mostly compared datasets from healthy young individuals with those from older patients, without age-matched controls, which may introduce bias. For example, Fung et al. reported a decrease in liver stiffness in a large Chinese population, from a median of 4.2 kPa in those aged under 25 years to 3.4 kPa in those aged at least 55 years (36). In contrast, Sharma et al. did not find an age-dependency in an Indian population in the Uttarakhand region (37). However, even if liver softening was present in older individuals, as described by Fung et al., the respective bias would not have impeded the findings of our study. Rather, the increased liver softness in older individuals would be expected to amplify the pulsation artifact, thereby enhancing the diagnostic performance of features such as *q1*. Furthermore, the patient examinations were conducted with diverse acquisition settings over a relatively long study period. From a puristic standpoint, this may be a limitation. However, it may suggest that our findings are generalizable. One problem might be that we included patient data acquired under both respiratory-triggered and free-breathing conditions. This may not be a major limitation, however, since the dependence of the cardiac pulsation artifact on the breathing cycle was reported to be small [38]. A further limitation is that the proposed method only assesses the left liver and is therewith blind to cirrhotic alterations of the right liver lobe. Finally, we only included first-order statistical features, whereas the inclusion of higher-order radiomic features may provide a more detailed quantification of artifacts in the left liver lobe. Incorporating such features could potentially improve the predictive power but would complicate the analysis and potentially reduce the clinical translation potential. The use of a single, visually well-discernable feature such as *q1*, however, might be more advantageous in a practical setting than the use of many advanced features.

## Conclusion

In conclusion, we demonstrated in a quantitative, retrospective analysis, that the DWI liver pulsation artifact is a potential marker for liver fibrosis assessment. Whether this approach is suitable for clinical practice, where qualitative image assessments are the norm, remains to be evaluated in a prospective clinical study. It could be valuable, particularly since DWI data are acquired in most liver MRI protocols, enabling the acquisition of additional information without prolonging the scan duration.

## Supporting information

raw Pixel values for left and right liver lobe of different b-values and patients

## Data Availability

The minimal data set including a explanation of the data is attached in the supplements (data.zip).

## Appendix

Figure 6 shows example images of a false-negative and a false-positive case.

**Figure 6:**
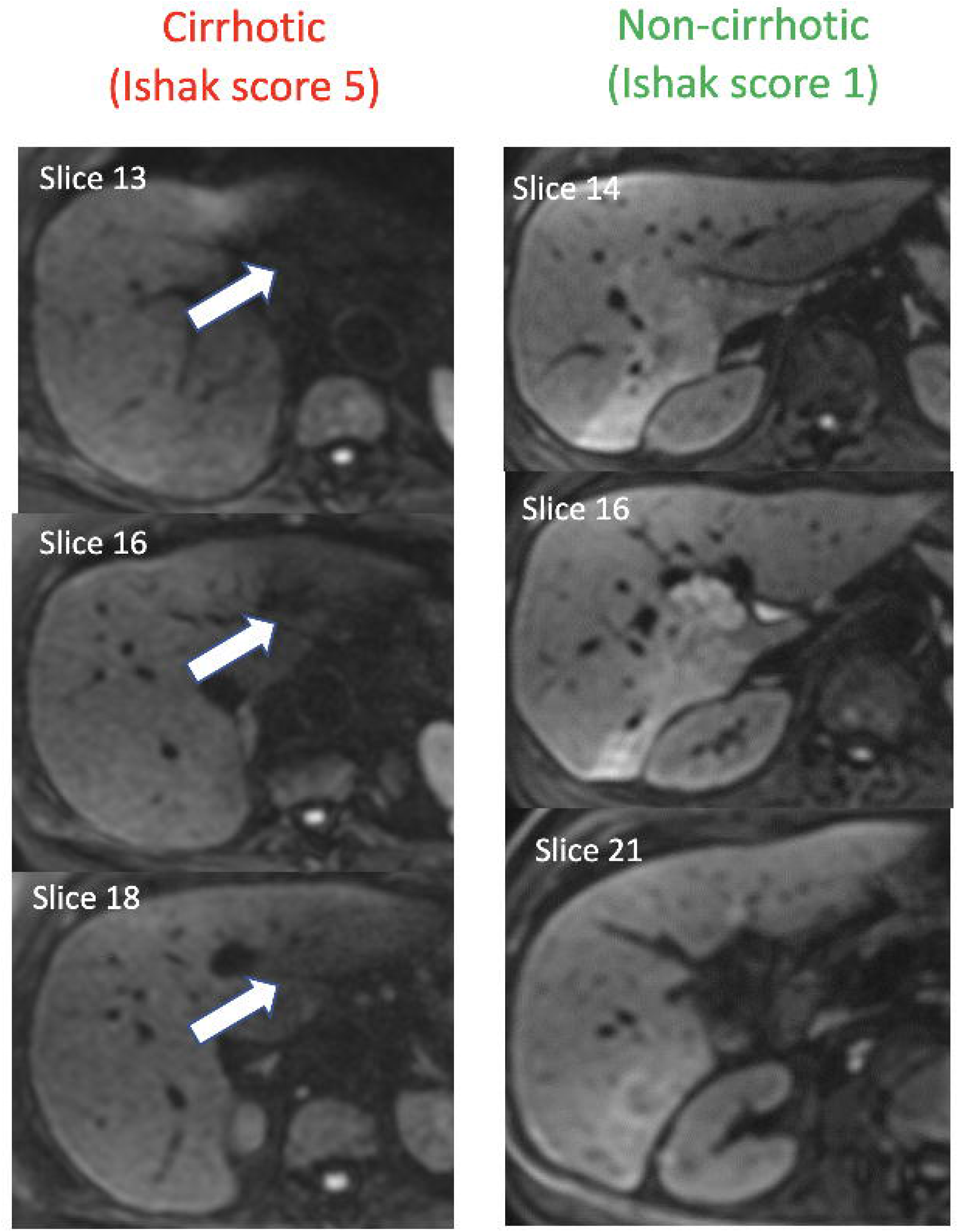
Representative examples of a false-negative (cirrhotic, Ishak score 5) and false-positive (non-cirrhotic, Ishak score 1) case. Each column shows three b = 800 s/mm² trace-weighted images of a cirrhotic or healthy liver. The left column shows a patient with cirrhosis and a visible artifact; the right column shows a patient with an Ishak score of 1 and an apparent artifact, indicated by white arrows.

## Acknowledgments

Retrieving retrospective imaging data and patient information is part of the MD thesis/project of Carolin Arnold. We thank Peter Dietrich from the department of internal medicine 1 for assisting with retrospective patient data selection. We thank Claus Hellerbrand for his support of this work.

## Abbreviations

DWI: diffusion-weighted imaging
MRI: Magnetic resonance imaging
ROI: Region of interest
EPI: echo planar imaging
GRAPPA: GeneRalized Autocalibrating Partial Parallel Acquisition
SMS: simultaneous multislice
MDDW: multi-directional diffusion weighting
SPAIR: Spectral Adiabatic Inversion Recover
AUC: Area Under Curve
ROC: Receiver Operator Characteristics
q1: 25^th^ percentile
p10: 10^th^ percentile
median: 50^th^ percentile
q3: 75^th^ percentile
p90: 90^th^ percentile
mad: mean absolute deviation
std: standard deviation
rms: root mean squared
rmad: robust mean absolute deviation
iqr: interquartile range

